# Glasses Against transmission of SARS-CoV-2 in the community (GLASSY): a pragmatic randomized trial

**DOI:** 10.1101/2022.07.31.22278223

**Authors:** Atle Fretheim, Ingeborg Hess Elgersma, Arnfinn Helleve, Petter Elstrøm, Oliver Kacelnik, Lars G. Hemkens

## Abstract

**Background:** Observational studies have reported an association between use of eye protection and reduced risk of SARS-CoV-2 infection and other respiratory viruses, but as for most non-pharmaceutical interventions for infection control, no randomized trials have been conducted. We conducted a randomized trial to evaluate the effectiveness of recommending the use of glasses in public as protection against being infected with SARS-CoV-2 and other respiratory viruses.

**Methods and findings:** This was a pragmatic, randomized, trial in Norway from 2 February to 24 April 2022 where all adult members of the public who did not regularly wear glasses, had no symptoms of COVID-19 and no COVID-19 in the last 6 weeks, were eligible. Participants randomized to the intervention group were asked to wear glasses (e.g. sunglasses) for 2 weeks when close to others in public spaces. The primary outcome was positive COVID-19 test result notified to the Norwegian Surveillance System for Communicable Diseases (MSIS). Secondary outcomes included positive COVID-19 test result based on self-report and episode of respiratory infection based on self-report of symptoms. We randomized 3717 participants. All were identified and followed up in the registries and 87% responded to the end of study-questionnaire. The proportions with a notified positive COVID-19 test in the national registry were 3.7% in the intervention group (68/1852) and 3.5% (65/1865) in the control group (95% CI for risk difference -1.0% to 1.4%; relative risk 1.10, 95% CI 0.75 to 1.50). The proportions with a positive COVID-19 test based on self-report were 9.6% and 11.5% (95% CI for risk difference -3.9% to 0.1%; relative risk 0.83, 95% CI 0.69 to 1.00). The risk of respiratory infections based on self-reported symptoms was lower in the intervention group (31% vs. 34%; 95% CI for risk difference -3.3% to -0.3%; relative risk 0.90, 95% CI 0.82 to 0.99).

**Conclusions:** Our results indicate that asking people to wear glasses may protect the public against respiratory infections, but the findings are not certain, and the study needs replication. Although the evidence is uncertain, and the effect probably modest at best, wearing glasses may be worth considering as one component in the infection control toolbox since it is a simple, low burden, and low-cost intervention, with few negative consequences.

**Trial registration:** ClinicalTrials.gov Identifier: <u>NCT05217797</u>

**Funding:** The costs of running the trial were covered by the Centre for Epidemic Interventions Research (CEIR), Norwegian Institute of Public Health. The authors received no specific funding for this work

## Introduction

Use of eye protective gear for infection control was proposed over 100 years ago [1], but has received little attention during the ongoing COVID-19 pandemic, except as part of Personal Protective Equipment (PPE) for health care workers [2].

A simple means of eye protection is to wear glasses. Many have easy access to sunglasses, and it requires little effort to use them in everyday life. Repurposing sunglasses for infection control could be a simple, readily available, environmentally friendly, safe, and sustainable infection prevention measure. An association between wearing glasses and lower risk of respiratory viruses has been reported from observational studies, but to our knowledge no randomized trial of eye protection against respiratory virus infection has been conducted, neither in health care nor in community settings [3-6].

To inform decisions about the use of personal protection against SARS-CoV-2, we carried out a randomized trial of wearing glasses to reduce the risk of being infected with SARS-CoV-2 and other respiratory viruses.

## Methods

The study protocol was published prior to the start of the study [7]. There were no major deviations from the protocol. We followed the CONSORT reporting guidelines for pragmatic trials [8].

### Participants

We recruited participants via an online portal that we distributed to the Norwegian public through the media, online and print adverts and e-mails to members of survey panels of two data collection companies. All members of the public could take part who

1. were at least 18 years of age
2. did not regularly wear glasses
3. owned or could borrow glasses that they could use (e.g. sun-glasses)
4. had not contracted COVID-19 in the six weeks prior to participation
5. did not have COVID-19 symptoms when providing consent
6. were willing to be randomized to wear, or not wear glasses outside their home when close to others for a 2-week period.
7. provided informed consent

Those dependent on visual aids who could use contact lenses if allocated to the control group, were eligible.

### Intervention

The participants allocated to the intervention group were asked to wear sunglasses or other types of glasses when close to other people outside their home (on public transport, in shopping malls etc.), over a 14-day period. The control group was encouraged not to wear glasses when close to others outside their home.

### Objective

We aimed to evaluate whether wearing glasses has an impact on the risk of being infected by the SARS-CoV-2 virus or developing a respiratory infection.

### Outcomes

All data were collected from national registries or the end of study survey. In general, data from administrative registries in Norway are reliable and of high quality and are used extensively in research [9]. All participants had to identify themselves using their personal ID-code, and were thereby identifiable in the national health registries. All PCR COVID-19 tests are analyzed by laboratories that report results directly to the Norwegian Surveillance System for Communicable Diseases (MSIS), so we can reasonably assume that all such tests are registered.

#### Primary outcome

- Any positive COVID-19 test result notified to the Norwegian Surveillance System for Communicable Diseases (MSIS), from day 3 to day 17 of the study period.

#### Secondary outcomes

- Any positive COVID-19 test result based on self-report, from day 1 to day 17 of the study period.
- Episode of respiratory infection based on self-report of symptoms from day 1 to day 17 of the study period. We defined respiratory infection as having one respiratory symptom (stuffed or runny nose, sore throat, cough, sneezing, heavy breathing) and fever, or one respiratory symptom and at least two more symptoms (body ache, muscular pain, fatigue, reduced appetite, stomach pain, headache, loss of smell,)
- Health care use for respiratory symptoms, self-reported, from day 1 to day 17 of the study period
- Health care use for injuries, self-reported, from day 1 to day 17 of the study period
- Health care use (all causes), self-reported, from day 1 to day 17 of the study period
- Health care use for respiratory symptoms as registered in Norwegian Patient Registry (NPR), from day 3 to day 28 of the study period
- Health care use for injuries (from day 1 to day 21 as registered in NPR and the Norwegian Registry for Primary Health Care (KPR), from day 3 to day 28 of the study period
- Health care use (all causes) as registered in NPR and KPR from day 1 to day 21 of the study period

We do not report on the latter four outcomes here as we do not have access to this data for the time being.

We also asked the participants about adherence to the intervention, use of face masks, testing behavior, and whether they had any negative experiences with taking part in the study. See supporting information (S1) for questionnaire.

### Sample size

We originally estimated a need for around 22 000 participants. This was based on an expected event rate of 2% in the control group and 1.5% in the intervention group. The underlying assumptions are provided in the study protocol [7].

### Randomization and masking

The participants were automatically randomized and informed about which group they were allocated to immediately after signing the consent form in the online recruitment platform (Nettskjema). No masking was feasible, except for in the analysis phase. One member of our team (AH) provided the chief analyst (IHE) with a data file where group allocation was blinded. The chief analyst presented the main results to the project team who were also blinded to the group allocation (and data on use of glasses). We discussed how we would interpret the findings depending on whether one group or the other was the intervention group, and we prepared a short report that we posted online before unblinding ourselves to the group allocation [10].

### Statistical methods

We conducted unadjusted analyses to estimate the relative risk and risk difference for the prespecified outcomes, with 95% confidence intervals calculated with the Wald method. All analyses adhered to the intention to treat principle and we included all randomized participants in the denominator in all our main analyses. Where outcome data was missing, we assumed that there was no event (no positive COVID-19 test, no symptoms, no use of health care services etc.). As a post hoc sensitivity analysis, we also conducted a crude (unadjusted) per protocol analysis where we excluded participants using glasses in the control group and participants not using glasses in the intervention group.

As planned, we conducted a subgroup analysis for participants that used contact lenses, with the hypothesis that the effect would be smaller for this group since control group participants were allowed to use contact lenses. One possible mechanism for viral transmission is that the viruses attach to ACE-2 receptors in the cornea, which we assume are protected by contact lenses [5]. We also conducted subgroup analyses for groups with different vaccination status and for those with a history of COVID-19. Differences in treatment effects across subgroups were assessed using a chi-square test of interaction.

The trial is registered at ClinicalTrials.gov (NCT05217797).

### Data management

We used the University of Oslo’s web-based survey platform Nettskjema for screening and data collection, and their service for secure storage of research data (TSD).

We collected directly identifiable data (name, person identification number and e-mail address). Along with a code for linking data, the person identification number was sent to the registries. Each register deleted the person identification number before register-data and code for linking were delivered and stored in TSD. The questionnaire data were processed in the same way. The codes for linking data to the person identifier was kept in a securely stored database, with limited access.

## Ethics

Information about the goal of the trial and eligibility study were provided through an online portal, and eligible participants consented to participation using the national digital signature service (Posten signering). In our judgement, taking part entailed negligible risk and the findings can potentially inform decisions about infection control measures in the ongoing and future epidemics. Thus, we considered the risk-benefit ratio morally defensible.

The protocol was approved by the Regional Ethics Committee of South-East Norway (application number 428685).

## Results

Recruitment commenced on 2 February 2022. Of the 7506 individuals who showed interest in participating in the trial by completing the screening form, 3717 were randomized (Figure 1). Most participants entered the trial soon after initiation, following wide media coverage of the launch of the trial on Norwegian national television and other media outlets (Figure 2). Another wave of recruitment occurred after two data collection companies invited members of their survey panels to take part in the trial. We closed recruitment 24 April 2022. Most participants were female, had a mean age of 47 years, and had received three or more doses of vaccination against COVID-19 (Table 1).

**Fig 1.**
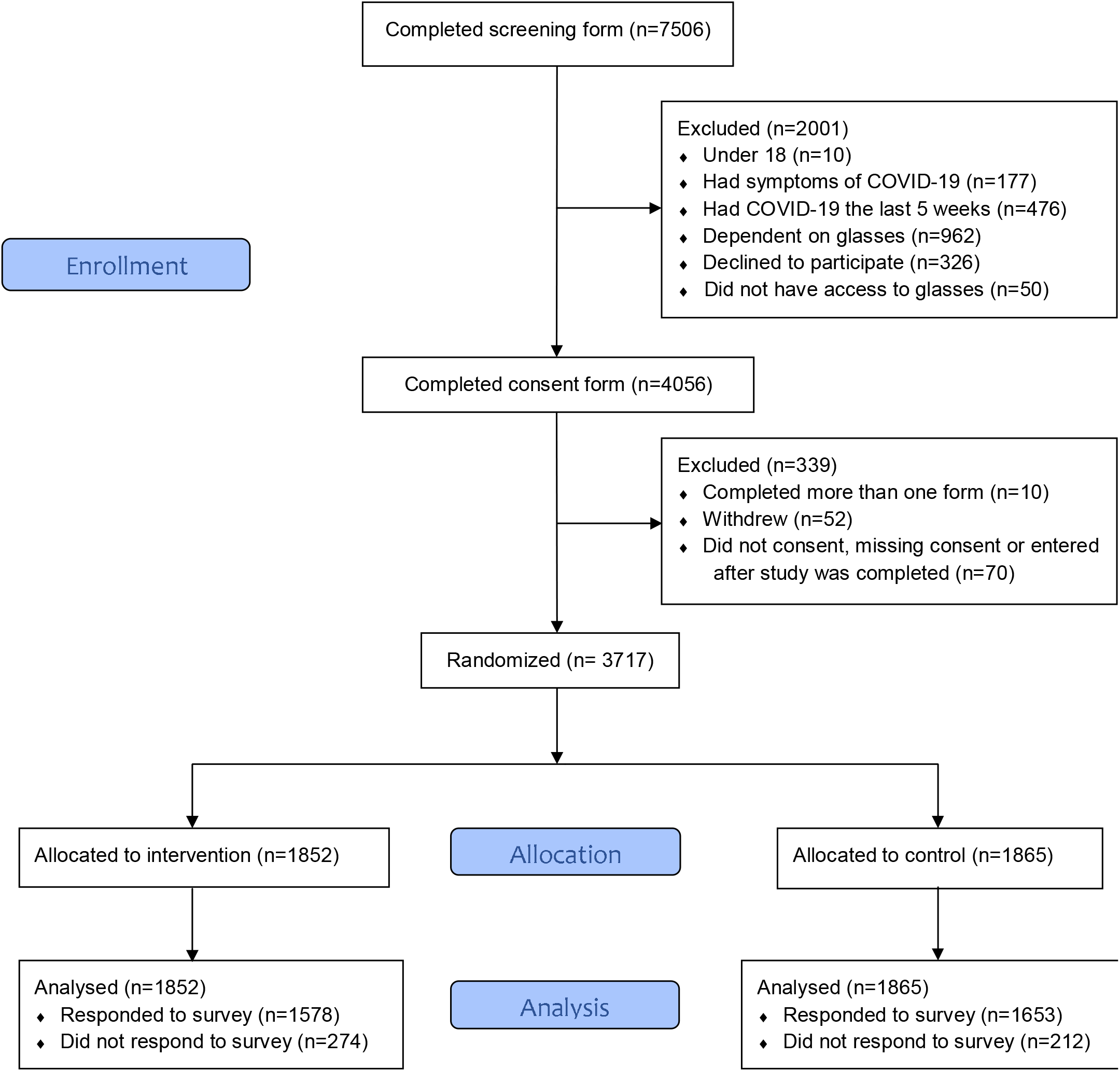
Flow diagram of participants.

**Fig 2.**
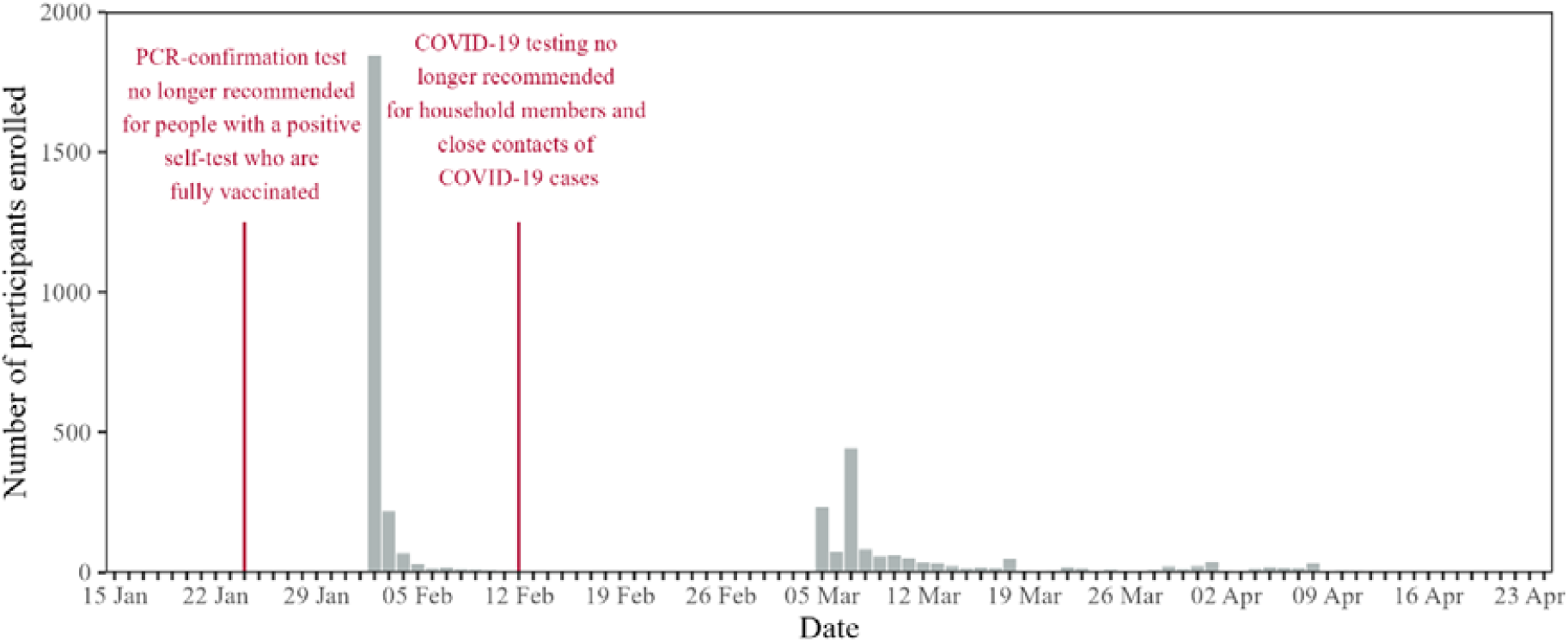
Recruitment over time. Grey bars represent the number of participants enrolled each day. Red vertical lines represent major changes in testing recommendations from national health authorities.

**Table 1.** Baseline characteristics of trial participants.

|  | Intervention group (n=1852) | Control group (n=1865) |
| --- | --- | --- |
| Female | 1193 (64.4%) | 1246 (66.8%) |
| Male | 659 (35.6%) | 619 (33.2%) |
| Mean age at inclusion (sd) | 46.9 (15.1) | 46.8 (15.1) |
| Number of COVID-19 vaccines received |  |  |
| 0 | 50 (2.7%) | 58 (3.1%) |
| 1 | 19 (1.0%) | 21 (1.1%) |
| 2 | 361 (19.5%) | 335 (18.0%) |
| 3 or more | 1422 (76.8%) | 1451 (77.8%) |

The main findings are summarized in Table 2. For notified COVID-19 cases the proportions were 68/1852 (3.7%) in the intervention group, and 65/1865 (3.5%) in the control group (RR 1.10; 95% CI 0.76 to 1.50). The proportion with self-reported positive tests was lower in the intervention group, 177/1852 (9.6%) vs. 214/1865 (11.3%) (RR 0.83; 95% CI 0.69 to 1.00). Almost a third of all participants reported symptoms of respiratory infection, with a lower proportion in the intervention group than in the control group (RR 0.90; 95% CI 0.82 to 0.99). There were few notified COVID-19 cases among the non-respondents to the survey (8 in the intervention group and 3 in the control group).

**Table 2.** Main findings.

|  | Intervention group<br>(n=1852) | Control group<br>(n=1865) | Relative<br>risk | Absolute<br>risk<br>difference |
| --- | --- | --- | --- | --- |
| Notified COVID-19 case<sup>1</sup> | 68/1852 (3.7%) | 65/1865 (3.5%) | 1.10 (0.75<br>to 1.50) | 0.2% (-1.0<br>to 1.4) |
| Self-reported COVID-19<br>case<sup>2</sup> | 177/1852 (9.6%) | 214/1865<br>(11.5%) | 0.83 (0.69<br>to 1.00) | -1.9% (-3.9<br>to 0.1) |
| Respiratory infection<sup>3</sup> | 571/1852 (30.8%) | 636/1865<br>(34.1%) | 0.90 (0.82<br>to 0.99) | -3.3% (-6.3<br>to -0.3) |
| Health care use, all cause | 80/1852 (4.3%) | 83/1865 (4.5%) | 0.97 (0.72<br>to 1.30) | -0.1% (-1.4<br>to 1.2) |
| - Health care use, due<br>to airway symptoms<sup>3</sup> | 15/1852 (0.8%) | 20/1865 (1.1%) | 0.76 (0.39<br>to 1.50) | -0.3% (-0.9<br>to 0.4) |
| - Health care use, due<br>to injuries<sup>3</sup> | 24/1852 (1.3%) | 16/1865 (0.9%) | 1.50 (0.81<br>to 2.80) | 0.4% (-0.2<br>to 1.1) |

<sup>1</sup> Date of test between day 3 and day 17 after inclusion in the study
<sup>2</sup> Date of test Between day 1 and day 17 after inclusion in the study
<sup>3</sup>*Self-reported*

We report behavioral outcomes in Table 3. In the intervention group, 71% of the participants reported using glasses often, almost always, or always, while the corresponding proportion in the control group was 11% (95% CI for difference 58 to 63%). The use of face masks was higher in the intervention-group (40% vs. 30%, 95% CI for difference 7 to 13%). The proportion of participants who took a COVID-test during the trial period was similar in the two groups (50% vs. 49%). We also asked the participants whether they normally commuted to work/studies by public transportation, and the proportion reporting that they did was lower in the intervention group (23% vs. 27%; 95% CI for difference -7 to -1%).

**Table 3.** Behavioral outcomes.

|  | Intervention group<br>(n=1852) | Control group<br>(n=1865) | Risk difference (95%<br>CIs) |
| --- | --- | --- | --- |
| Responded to survey | 1578/1852 (85.2%) | 1653/1865 (88.6%) | -3.4% (-5.6 to -1.3) |
| Use of glasses |  |  |  |
| At least 50% of the time | 1306/1852 (70.5%) | 196/1865 (10.5%) | 60.0% (57.5 to 62.5) |
| - Always | 575/1852 (31.0%) | 60/1865 (3.2%) | 27.8% (25.6 to 30.1) |
| - Almost always<br>(75 to 100% of<br>the time) | 524/1852 (28.3%) | 81/1865 (4.3%) | 24.0% (21.7 to 26.2) |
| - Often (50 to<br>75% of the<br>time) | 207/1852 (11.2%) | 55/1865 (2.9%) | 8.2% (6.6 to 9.9) |
| - Sometimes (25<br>to 50% of the<br>time) | 91/1852 (4.9%) | 59/1865 (3.2%) | 1.8% (0.5 to 3.0) |
| - Almost never<br>(up to 25% of<br>the time) | 101/1852 (5.5%) | 339/1865 (18.2%) | -12.7% (-14.8 to -10.7) |
| - Never | 78/1852 (4.2%) | 1054/1865 (56.5%) | -52.3% (-54.7 to -49.9) |
| Use of facemasks |  |  |  |
| At least 50% of the time | 731/1852 (39.5%) | 554/1865 (29.7%) | 9.8% (6.7 to 12.8) |
| - Always | 180/1852 (9.7%) | 134/1865 (7.2%) | 2.5% (0.7 to 4.3) |
| - Almost always<br>(75 to 100% of<br>the time) | 319/1852 (17.2%) | 225/1865 (12.1%) | 5.2% (2.9 to 7.4) |
| - Often (50 to<br>75% of the<br>time) | 232/1852 (12.5%) | 195/1865 (10.5%) | 2.1% (0.0 to 4.1) |
| - Sometimes (25<br>to 50% of the<br>time) | 210/1852 (11.3%) | 227/1865 (12.2%) | -0.8% (-2.9 to 1.2) |
| - Almost never<br>(up to 25% of<br>the time) | 250/1852 (13.5%) | 345/1865 (18.5%) | -5.0% (-7.4 to -2.6) |
| - Never | 381/1852 (20.6%) | 524/1865 (28.1%) | -7.5% (-10.3 to -4.8) |
| Use of COVID-19 tests |  |  |  |
| Tested | 925/1852 (49.9%) | 914/1865 (49.0%) | 0.9% (-2.3 to 4.2) |
| - Home test and<br>test station | 120/1852 (6.5%) | 103/1865 (5.5%) | 1.0% (-0.6 to 2.5) |
| - Only test station | 10/1852 (0.5%) | 13/1865 (0.7%) | -0.2% (-0.7 to 0.3) |
| - Only home test | 795/1852 (42.9%) | 798/1865 (42.8%) | 0.1% (-3.0 to 3.3) |
| - No | 651/1852 (35.2%) | 739/1865 (39.6%) | -4.5% (-7.6 to -1.4) |

The results of the per protocol analysis, i.e. where we excluded non-adherent participants in both groups, are presented in Table S1. There were practically no differences between these estimates and those of the main analysis. Subgroup analyses did not suggest any interaction effects, except between vaccination status and relative risk of notified COVID-19 (Tables S2 and S3).

In total 76 participants reported having negative experiences with taking part in the trial (53 in the intervention group and 23 in the control group), and all but one elaborated using free text. The most common complaint concerned the combination of glasses and face masks, especially fogging (21 in the intervention group mentioned this). Wearing glasses felt uncomfortable or tiring by some, and a few complained over reduced vision due to using sunglasses or reading glasses. One person in the intervention group reported a fall due to reduced vision. A few felt silly when wearing glasses or sunglasses, e.g. inside a cafe. In the control group there were some who reported headaches or other problems from not being able to use glasses.

## Discussion

The findings from this first randomized trial of eye protection against viral spread indicate that using glasses in public may protect against respiratory viruses, but the evidence is uncertain, and more trials are needed to corroborate this evidence.

### Comparison with other studies

A recent systematic review of studies estimating the impact of eye protection on transmission of SARS-CoV-2 identified 5 observational studies, all conducted among health workers in various settings [5]. With one exception, all studies reported a clear association between use of eye protection and reduced infection risk, with estimated relative risk reductions ranging from 40% to 96%. The systematic review authors also identified three cross sectional studies examining the use of glasses in the community, all of which found a substantial underrepresentation of people wearing glasses among COVID-19 patients [11-13]. In addition, some studies have shown high detection rates of SARS-CoV-2 in ocular fluids from COVID-19 patients, also indicating that the eyes may be an important route of viral transmission [14, 15]. An earlier systematic review, from 2020, of studies from previous coronavirus-outbreaks (MERS and SARS), included 12 studies in a meta-analysis indicating that use of eye protection was associated with considerable reduced infection risk (relative risk 0.34; 95% CI 0.21 to 0.56) [4]. None of the studies were randomized, and nearly all were conducted in health care settings. Furthermore, a Cochrane-review from 2020 identified no randomized trials of eye protection to curb the spread of respiratory viruses [3]. While we were conducting our trial, a pre-print with findings from a cohort study in the UK was published, reporting 15% lower odds of COVID-19 among consistent users of glasses (OR 0.85, 95% 0.77 to 0.94), after adjusting for key variables e.g. age [16]. Overall, all these prior findings are compatible with the results of this first large randomized trial.

Our trial is one of very few randomized trials evaluating public health and social measures for epidemic control [3, 4, 6]. For example, only two trials of face masks have been conducted during the COVID-19 pandemic [17, 18]. The results from the two face mask trials are, broadly speaking, in the same range as ours, i.e. consistent with a modest albeit uncertain effect.

### Strengths and limitations of study

The result for our primary endpoint is very uncertain, with a wide confidence interval compatible with both important benefit and harm. If we only emphasize this outcome, our trial can be interpreted as largely “negative” although the confidence interval around the estimate includes a 25% relative reduction, which is what we had deemed a reasonable minimal important difference in our sample size estimation [7]. We had probably achieved a more precise estimate had we been more successful with our recruitment of participants. Despite wide media coverage, we came nowhere near our target of 22000. Also, a week before we opened recruitment into the trial, the health authorities changed its recommendations for testing, no longer advising a confirmatory PCR-test for fully vaccinated people who self-tested positive for COVID-19. Then, 10 days after we started recruitment, the authorities changed its testing recommendations again, revoking the earlier advise that members of households with a COVID-19-case should get tested, and recommended that only adults with symptoms should test themselves. These policy changes were followed by a drastic reduction in the number of test results reported to the Norwegian Surveillance System for Communicable Diseases (MSIS), i.e. the data source for our main outcome. We therefore considered changing our main outcome from notified COVID-19 cases to self-reported cases but decided to adhere to the protocol. In our interpretation of the study findings, we have put more weight on the secondary outcomes, i.e. self-reported positive COVID-19 test and symptoms consistent with respiratory infection. This decision can reasonably be challenged.

There is risk of bias for our survey-based outcomes, since we lack responses from 13% of the participants. The fact that there were very few notified COVID-19 cases among the non-respondents indicates that this risk is low.

We believe the pragmatic trial approach we adopted is particularly appropriate for evaluations of public health interventions where real life effectiveness is of key interest, and more so than being able to demonstrate effects under ideal conditions (e.g. in a laboratory), or to understand underlying mechanisms [19-21]. We are not able to clearly decipher to what extent the observed effects were due to increased wearing of glasses, increased use of face masks, differences in the use of public transport, decreased awareness of symptoms or decreased reporting of outcomes. Consistent with the pragmatic intent, we sought to be as close as possible to the natural environment and avoid artificial research scenarios that burden participants with data collection beyond a short questionnaire at the end of the study. However, we collected some specific information to better understand how the intervention worked, and this is consistent with the hypothesized mechanism that eye protection contributes to reduced transmission of respiratory infections. One of the items in the questionnaire concerned the participants’ use of public transport for their daily commute and was meant to capture their use of public transport at baseline. Interpreting the responses is not straightforward since the data were collected later, so the responses may have been influenced by taking part in the trial, including exposure to the intervention.

A surprise finding to us is that the use of face masks was substantially higher in the intervention group, especially since difficulties with combining glasses and face masks was the most common complaint among those who reported negative experiences.

While we believe that our findings are likely to be applicable in many settings beyond ours, there are some factors that may reduce the applicability. First, we conducted our trial in a phase of the pandemic where the omicron variant of the virus dominated. It is possible that new variants, or other respiratory viruses, have different modes of transmission. Second, the trial took place during the Norwegian winter, and the importance of transmission in public spaces may be different in other seasons or cultural settings. Third, the COVID-19 incidence was exceptionally high during the time of the trial. Fourth, the effects that the intervention had on behavior, e.g. face mask use, will likely vary across contexts and populations.

## Conclusions

The results from our trial seem to support our hypothesis, i.e. that recommending use of glasses in public reduces the risk of being infected by respiratory viruses. The evidence is uncertain, and the effect probably modest at best, but since the intervention is simple, low burden, low-cost, and with few negative consequences, it is worth considering as one component in the infection control toolbox. We urge others to conduct similar simple, pragmatic trials.

## Supporting information

Supplementary file 1. Questionnaire

## Data Availability

All data produced in the present study are available upon reasonable request to the authors.

## Declarations

### Authors’ contributions

AF was the Chief Investigator and led the protocol development. LGH had the original idea for the study, presenting it in September 2020. AH was responsible for the data collection platform. IHE was the chief analyst and conducted all statistical analyses, with support from PE who also reviewed the data and the analyses. AF and IHE wrote the first manuscript draft. All authors contributed to study design and development of the protocol, and all authors commented on and approved the final version of the manuscript.

### Funding

All running costs were covered by the Centre for Epidemic Interventions Research (CEIR), Norwegian Institute of Public Health.

### Competing interests

All authors declare: no competing interests.

## Acknowledgements

Christopher James Rose helped with the statistical analyses. We thank journalist Hallvard Sandberg who posted a tweet on 4 January 2022, challenging us to investigate the potential of using glasses as protection against transmission. Dagfinn Bergsager (University of Oslo) facilitated our use of the web-based questionnaire and safe data storage. For data storage and analyses we used the TSD (Tjeneste for Sensitive Data) facilities, owned by the University of Oslo, operated and developed by the TSD service group at the University of Oslo, IT-Department (USIT). Finally, we thank all those who volunteered to take part in the trial.

## Data sharing statement

The final anonymized trial dataset will be freely available to the public.

---

**Table S1. Crude per protocol analysis**, i.e. only including those in the intervention group who reported wearing glasses more than 50% of the time, and those in the control group who reported wearing glasses less than 50% of the time.

**Table S2. Subgroup analysis using notified COVID-19 cases as outcome**.

**Table S3. Subgroup analysis for using self-reported COVID-19 cases as outcome**.

## Notes

### Competing Interest Statement

The authors have declared no competing interest.

### Clinical Trial

NCT05217797

### Clinical Protocols

https://www.medrxiv.org/content/10.1101/2022.02.04.22270120v1

https://zenodo.org/record/6521815#.YuT1PHZByUk

https://zenodo.org/record/6669688#.YuT1UnZByUk

### Funding Statement

This study did not receive any funding.

### Author Declarations

The Regional Ethics Committee (REK) of South-East Norway gave ethical approval for this work (application number 428685).

## References

1. Maxcy KF. The Transmission of infection through the eye. Journal of the American Medical Association. 1919;72(9):636–9. doi: 10.1001/jama.1919.02610090020005.

2. Coroneo MT, Collignon PJ. SARS-CoV-2: eye protection might be the missing key. Lancet Microbe. 2021;2(5):e173–e4. Epub 2021/03/04. doi: 10.1016/S2666-5247(21)00040-9. PubMed PMID: 33655228; PubMed Central PMCID: PMCPMC7906687.

3. Jefferson T, Del Mar CB, Dooley L, Ferroni E, Al-Ansary LA, Bawazeer GA, et al. Physical interventions to interrupt or reduce the spread of respiratory viruses. Cochrane Database Syst Rev. 2020;11:CD006207. Epub 2020/11/21. doi: 10.1002/14651858.CD006207.pub5. PubMed PMID: 33215698; PubMed Central PMCID: PMCPMC8094623.

4. Chu DK, Akl EA, Duda S, Solo K, Yaacoub S, Schunemann HJ, et al. Physical distancing, face masks, and eye protection to prevent person-to-person transmission of SARS-CoV-2 and COVID-19: a systematic review and meta-analysis. Lancet. 2020;395(10242):1973–87. Epub 2020/06/05. doi: 10.1016/S0140-6736(20)31142-9. PubMed PMID: 32497510; PubMed Central PMCID: PMCPMC7263814.

5. Byambasuren O, Beller E, Clark J, Collignon P, Glasziou P. The effect of eye protection on SARS-CoV-2 transmission: a systematic review. Antimicrob Resist Infect Control. 2021;10(1):156. Epub 2021/11/06. doi: 10.1186/s13756-021-01025-3. PubMed PMID: 34736533; PubMed Central PMCID: PMCPMC8567128.

6. Hirt J, Janiaud P, Hemkens LG. Randomized trials on non-pharmaceutical interventions for COVID-19: a scoping review. BMJ Evid Based Med. 2022. Epub 2022/01/29. doi: 10.1136/bmjebm-2021-111825. PubMed PMID: 35086864; PubMed Central PMCID: PMCPMC8804305.

7. Fretheim A, Hemkens LG, Helleve A, Elgersma IH, Elstrøm P, Kacelnik O. The GLasses Against transmission of SARS-CoV-2 in the communitY (GLASSY) trial: A pragmatic randomized trial (study protocol). medRxiv. 2022:2022.02.04.22270120. doi: 10.1101/2022.02.04.22270120.

8. Zwarenstein M, Treweek S, Gagnier JJ, Altman DG, Tunis S, Haynes B, et al. Improving the reporting of pragmatic trials: an extension of the CONSORT statement. BMJ. 2008;337:a2390. Epub 2008/11/13. doi: 10.1136/bmj.a2390. PubMed PMID: 19001484; PubMed Central PMCID: PMCPMC3266844.

9. Lovlie A, Blystad H, Bruun T. MSIS celebrates 40 years. Tidsskr Nor Laegeforen. 2015;135(23-24):2136–8. Epub 2015/12/18. doi: 10.4045/tidsskr.15.1028. PubMed PMID: 26674030.

10. Blinded assessment of GLASSY-study results [Internet]. Zenodo. 2022 [cited 24.06.2022].

11. Lehrer S, Rheinstein P. Eyeglasses Reduce Risk of COVID-19 Infection. In Vivo. 2021;35(3):1581–2. Epub 2021/04/30. doi: 10.21873/invivo.12414. PubMed PMID: 33910839; PubMed Central PMCID: PMCPMC8193301.

12. Saxena AK. Risk of COVID-19 among Spectacles Wearing Population of Northern India. Journal of Clinical and Diagnostic Research. 2021;15(5):NC08–NC11. Epub May 1,2021. doi: 10.7860/jcdr/2021/48079.14899.

13. Zeng W, Wang X, Li J, Yang Y, Qiu X, Song P, et al. Association of Daily Wear of Eyeglasses With Susceptibility to Coronavirus Disease 2019 Infection. JAMA Ophthalmol. 2020;138(11):1196–9. Epub 2020/09/17. doi: 10.1001/jamaophthalmol.2020.3906. PubMed PMID: 32936214; PubMed Central PMCID: PMCPMC7495310.

14. Azzolini C, Donati S, Premi E, Baj A, Siracusa C, Genoni A, et al. SARS-CoV-2 on Ocular Surfaces in a Cohort of Patients With COVID-19 From the Lombardy Region, Italy. JAMA Ophthalmol. 2021;139(9):956–63. Epub 2021/03/05. doi: 10.1001/jamaophthalmol.2020.5464. PubMed PMID: 33662099; PubMed Central PMCID: PMCPMC7934077.

15. Zhang X, Chen L, Wang G, Chen L, Huang L, Cao Y, et al. Investigation of SARS-CoV-2 on Ocular Surface of Coronavirus Disease 2019 Patients Using One-Step Reverse-Transcription Droplet Digital PCR. Infect Drug Resist. 2021;14:5395–401. Epub 2021/12/24. doi: 10.2147/IDR.S335635. PubMed PMID: 34938087; PubMed Central PMCID: PMCPMC8685385.

16. Navaratnam AMD, O’Callaghan C, Beale S, Nguyen V, Aryee A, Braithwaite I, et al. Glasses and risk of COVID-19 transmission - analysis of the Virus Watch Community Cohort study. medRxiv. 2022:2022.03.29.22272997. doi: 10.1101/2022.03.29.22272997.

17. Abaluck J, Kwong LH, Styczynski A, Haque A, Kabir MA, Bates-Jefferys E, et al. Impact of community masking on COVID-19: A cluster-randomized trial in Bangladesh. Science. 2022;375(6577):eabi9069. Epub 2021/12/03. doi: 10.1126/science.abi9069. PubMed PMID: 34855513; PubMed Central PMCID: PMCPMC9036942.

18. Bundgaard H, Bundgaard JS, Raaschou-Pedersen DET, von Buchwald C, Todsen T, Norsk JB, et al. Effectiveness of Adding a Mask Recommendation to Other Public Health Measures to Prevent SARS-CoV-2 Infection in Danish Mask Wearers : A Randomized Controlled Trial. Ann Intern Med. 2021;174(3):335–43. Epub 2020/11/19. doi: 10.7326/M20-6817. PubMed PMID: 33205991; PubMed Central PMCID: PMCPMC7707213.

19. Schwartz D, Lellouch J. Explanatory and pragmatic attitudes in therapeutical trials. J Chronic Dis. 1967;20(8):637–48. Epub 1967/08/01. doi: 10.1016/0021-9681(67)90041-0. PubMed PMID: 4860352.

20. Treweek S, Zwarenstein M. Making trials matter: pragmatic and explanatory trials and the problem of applicability. Trials. 2009;10:37. Epub 2009/06/06. doi: 10.1186/1745-6215-10-37. PubMed PMID: 19493350; PubMed Central PMCID: PMCPMC2700087.

21. Zwarenstein M. ‘Pragmatic’ and ‘explanatory’ attitudes to randomised trials. J R Soc Med. 2017;110(5):208–18. Epub 2017/05/16. doi: 10.1177/0141076817706303. PubMed PMID: 28504072; PubMed Central PMCID: PMCPMC5438067.

