## Supplementary file 1. Questionnaire for "Glasses Against transmission of SARS-CoV-2 in the community (GLASSY): a pragmatic randomized trial"

### S1 Questionnaire (translated from Norwegian)

| **Questions** | **Type of response** | **Options** |
| --- | --- | --- |
| 1. Do you wear contact lenses? | Categories | Yes, /No |
| 2. How often over the last two weeks have you been using glasses (reading glasses, sunglasses, sports glasses etc.) when you have been close to others outside your home? |  | Always/Almost always (at least 75% of the time)/ Often (50-75% of the time)/Sometimes (25-50 % of the time)/A few times (up to 25% of the time)/Never (0% of the time) |
| 3. How often over the last two weeks have you used a face mask when you have been close to others outside your home? |  | Always/Almost always (at least 75% of the time)/ Often (50-75% of the time)/Sometimes (25-50 % of the time)/A few times (up to 25% of the time)/Never (0% of the time) |
| 3. Do you normally use public transportation (bus, train, tram, metro, ferry, taxi) to and from work/study place? | Categories | Yes/No |
| 4. Have you experienced one of the following symptoms since taking part in the study? | Categories | Headache/Fever/Clogged nose or runny nose/ Reduced sense of smell/Reduced appetite/ Sore throat /Cough/ Sneezing/Body aches/ Muscle pain/Fatigue, lethargy/Heavy breathing/Abdominal pain |
| 5.Have you taken a COVID-19 over the last two weeks (self-test/test at testing facility)? | Categories | Yes, both self-test and at testing facility/Yes, but only a test station/Yes, but only self-test/No |
| 7. What was the test result? | Categories | Positive/Negative/Unsure |
| 8. When did you take the test? |  | Date |
| 9. Have you needed any medical care after you took part in the study? | Categories | Yes/No |
| Item 10-11 if response “Yes” on item 9 | | |
| 10. Did you need for medical care due to respiratory symptoms? | Categories | Yes/No |
| 11. Did you need for medical care due to injuries? | Categories | Yes/No |
| 12. Have you had any negative experiences from participating in this study? | Categories | Yes/No |
| Item 13 if response “Yes” on item 12 | | |
| 13. Please describe what those experiences were |  | Free text |
