## Supplementary file 2. Tables S1-S3 for "Glasses Against transmission of SARS-CoV-2 in the community (GLASSY): a pragmatic randomized trial"

### S2 Tables S1-S3

**Table S1. Crude per protocol analysis**, i.e. only including those in the intervention group who reported wearing glasses more than 50% of the time, and those in the control group who reported wearing glasses less than 50% of the time.

|  | Not wearing glasses in control group (n=1507) | Wearing glasses in intervention group (n=1306) | Risk ratio | Absolute risk difference |
| --- | --- | --- | --- | --- |
| Notified COVID-19 case¹ | 57/1507 (3.8%) | 50/1306 (3.8%) | 1.00 (0.70 to 1.50) | 0.0% (-1.4 to 1.5) |
| Self-reported COVID-19 case² | 201/1507 (13.3%) | 143/1306 (10.9%) | 0.82 (0.67 to 1.00) | -2.4% (-4.8 to 0.0) |
| Respiratory infection³ | 593/1507 (39.3%) | 462/1306 (35.4%) | 0.90 (0.82 to 0.99) | -4.0% (-7.6 to -0.4) |
| Health care use, all cause³ | 77/1507 (5.1%) | 69/1306 (5.3%) | 1.00 (0.75 to 1.40) | 0.2% (-1.5 to 1.8) |
| Health care use, due to airway symptoms³ | 16/1507 (1.1%) | 13/1306 (1.0%) | 0.94 (0.45 to 1.90) | -0.1% (-0.8 to 0.7) |
| Health care use, due to injuries³ | 15/1507 (1.0%) | 22/1306 (1.7%) | 1.70 (0.88 to 3.20) | 0.7% (-0.2 to 1.5) |

^1^Between day 3 and day 17 after inclusion in the study

^2^Between day 1 and day 17 after inclusion in the study

^3^Self reported

### Table S2. Subgroup analysis using notified COVID-19 cases as outcome.

| Variable | Subgroup | Intervention group | Control group | Relative risk | Absolute risk difference | P-value for interaction |
| --- | --- | --- | --- | --- | --- | --- |
| Lenses | Yes | 14/333 (4.2%) | 20/384 (5.2%) | 0.81 (0.41 to 1.60) | -1.0% (-4.1 to 2.1) | 0.41 |
|  | No | 46/1241 (3.7%) | 42/1267 (3.3%) | 1.10 (0.74 to 1.70) | 0.4% (-1.0 to 1.8) |  |
| Vaccine status | 0 | 5/50 (10.0%) | 4/58 (6.9%) | 1.40 (0.41 to 5.10) | 3.1% (-7.5 to 13.7) | 0.015 |
|  | 1 | 5/19 (26.3%) | 0/21 (0.0%) | - | 26.3% (6.5 to 46.1) |  |
|  | 2 | 23/361 (6.4%) | 30/335 (9.0%) | 0.71 (0.42 to 1.20) | -2.6% (-6.5 to 1.4) |  |
|  | 3+ | 35/1422 (2.5%) | 31/1451 (2.1%) | 1.20 (0.71 to 1.90) | 0.3% (-0.8 to 1.4) |  |
| COVID-19 previously | No | 66/1708 (3.9%) | 63/1743 (3.6%) | 1.10 (0.76 to 1.50) | 0.2% (-1.0 to 1.5) | 0.82 |
|  | Yes | 2/144 (1.4%) | 2/122 (1.6%) | 0.85 (0.12 to 5.90) | -0.3% (-3.2 to 2.7) |  |

### Table S3. Subgroup analysis for using self-reported COVID-19 cases as outcome.

| Variable | Subgroup | Intervention group | Control group | Relative risk | Absolute risk difference | P-value for interaction |
| --- | --- | --- | --- | --- | --- | --- |
| Lenses | Yes | 41/333 (12.3%) | 65/384 (16.9%) | 0.73 (0.51 to 1.00) | -4.6% (-9.8 to 0.5) | 0.26 |
|  | No | 135/1241 (10.9%) | 149/1267 (11.8%) | 0.93 (0.74 to 1.20) | -0.9% (-3.4 to 1.6) |  |
| Vaccine status | 0 | 6/50 (12.0%) | 6/58 (10.3%) | 1.20 (0.40 to 3.40) | 1.7% (-10.3 to 13.6) | 0.15 |
|  | 1 | 4/19 (21.1%) | 2/21 (9.5%) | 2.20 (0.46 to 11.00) | 11.5% (-10.7 to 33.7) |  |
|  | 2 | 39/361 (10.8%) | 60/335 (17.9%) | 0.60 (0.41 to 0.88) | -7.1% (-12.3 to -1.9) |  |
|  | 3+ | 128/1422 (9.0%) | 146/1451 (10.1%) | 0.89 (0.71 to 1.10) | -1.1% (-3.2 to 1.1) |  |
| COVID-19 previously | No | 175/1708 (10.2%) | 211/1743 (12.1%) | 0.85 (0.70 to 1.00) | -1.9% (-4.0 to 0.2) | 0.65 |
|  | Yes | 2/144 (1.4%) | 3/122 (2.5%) | 0.56 (0.096 to 3.30) | -1.1% (-4.4 to 2.3) |  |
